# Exploration of attitudes regarding uptake of COVID-19 vaccines among vaccine hesitant adults in the UK: A qualitative analysis

**DOI:** 10.1101/2021.12.23.21268325

**Authors:** Sarah Denford, Fiona Mowbray, Lauren Towler, Helena Wehling, Gemma Lasseter, Richard Amlôt, Isabel Oliver, Lucy Yardley, Mathew Hickman

## Abstract

**Background:** The aim of this work was to explore barriers and facilitators to uptake of COVID-19 vaccines and to explore views and reactions to efforts to improve vaccine uptake among those who were vaccine hesitant.

**Methods:** Semi-structured interviews were conducted with people between the age of 18-29 years who had not had a COVID-19 vaccine, and those between 30-49 years who had not received a second dose of a COVID-19 vaccine (more than 12 weeks after receiving a first).

**Results:** A total of 70 participants took part in the study, 35 participants had received one dose of the vaccine, and 35 had not received any vaccine.

Participants described a possible willingness to be vaccinated to keep themselves and those around them safe, and to avoid restrictions and return to normal. Barriers to uptake included: 1) perceived lack of need for COVID-19 vaccinations, 2) concerns about the efficacy of vaccinations, 3) concerns about safety, and 4) access issues. Uptake appeared to be influenced by the age and health status of the individual, trust in government and knowledge and understanding of science. Introduction of vaccine passes may provide a motive for having a vaccine but may also be viewed as coercive.

**Conclusion:** Participants were hesitant, rather than opposed, and had questions about their need for, and the safety and efficacy of the vaccine. Young people did not consider themselves to be at risk of becoming ill from COVID-19, did not think the vaccination was effective in preventing infection and transmission, and did not think sufficient research had been conducted with regard to the possible long-term side-effects. These concerns were exacerbated by a lack of trust in the government, and misunderstanding of science. In order to promote uptake, public health campaigns should focus on the provision of information from trusted sources that carefully explains the benefits of vaccination and addresses safety concerns more effectively. To overcome inertia in people with low levels of motivation to be vaccinated, appointments must be easily accessible.

## Introduction

The COVID-19 pandemic remains a substantial threat to the health and wellbeing of the UK population. The first vaccination against COVID-19 was approved in December 2020 and mass vaccination efforts are underway around the world. Globally, there are now more than 155 vaccine candidates, 495 ongoing vaccine trials being conducted in 61 countries, and 24 vaccines that have been approved for use [1]. Currently, individuals in the UK are offered one of three vaccines to protect against COVID-19; BNT162b2 mRNA (BioNTech, Pfizer vaccine), ChAdOx1 nCoV-19 (Oxford, AstraZenica vaccine), and mRNA-1273 SARS-CoV-2 (Moderna vaccine). In most cases two doses of the vaccine are required, given between 8 and 12 weeks apart, and a third “booster” dose is now being offered to all adults over the age of 18 years [2]. Prior to the identification of Omicron, the evidence showed that the efficacy of the vaccines was similar [3, 4]. However, following reports of a rare blood clotting problem linked to the AstraZeneca vaccine, it is no longer being offered to those under the age of 40 years [5]. The rollout of the COVID-19 vaccination program has been very successful in the UK so far, and as of November 2021, over 50 million people have received at least one vaccine and over 46 million people have received a second dose [6]. As emerging evidence highlights the importance of three doses for protection against variants of concern such as Omicron [7], and there is still low vaccine uptake among some groups [8, 9], efforts are urgently needed to support uptake.

Vaccine hesitancy refers to a delay or refusal of offered vaccinations [10]. Vaccine hesitant individuals may be uncertain or ambivalent about vaccination, but, with appropriate and effective public health messaging may be supported to accept vaccines in the future. A number of frameworks have been developed to describe individual level determinants for vaccine hesitancy: confidence, complacency, convenience (or constraints) [10], context and communication [11]. Research suggests high hesitancy regarding COVID-19 vaccines among certain groups [12]; such as those under the age of 50 years [12], those from minority ethnic groups [13], and those from most deprived areas [14]. There is some evidence to suggest that vaccine hesitancy is also slightly higher among women than men [13]. Understanding hesitancy among these groups is critical for supporting uptake of vaccinations in the future.

Although qualitative work has been conducted to explore attitudes towards vaccinations [15-18], much of that work was completed relatively early on in the pandemic during a time at which many groups hadn’t been offered the vaccine, and trials exploring the real world safety and efficacy of the vaccine were ongoing. As very little research has specifically explored attitudes regarding the second or third dose of the vaccine or age specific concerns for hesitant groups, the aim of this work was to explore attitudes, motives for and barriers to uptake of COVID-19 vaccines, to initiate discussions regarding how uptake can be improved, and to explore views and reactions to efforts to improve vaccine uptake among people under the age of 50 who had not received two doses of a COVID-19 vaccine.

## Methods

### Design

Interviews with people between the age of 18-29 years who had not had a COVID-19 vaccine, and those between 30-49 years who had not had a second dose of a COVID-19 vaccine more than 12 weeks after receiving their first dose.

### Sampling and recruitment

Participants were eligible for inclusion if they were: 1) currently living in the UK, 2) between the ages of 18 and 29 years and not had a COVID-19 vaccine, or 3) between 30 and 49 years and not had a second dose >12 weeks after the first dose, or did not intend to have a second dose of the vaccine.

There were two main routes for recruitment. First, we used social media (Facebook, Twitter and Instagram) to target participants living in areas in the UK in which uptake was low [19]. Study advertisements, shared through groups covering key areas in the UK with low uptake, encouraged unvaccinated individuals to register interest via an online sign up page (hosted by Qualtrics). We later commissioned a market research company (M3 Global Research) to share the study advertisement with eligible individuals on our behalf. Interested individuals were asked to leave basic demographic and contact details. We then used a purposive sampling strategy that aimed for diversity in age, gender, ethnic and socioeconomic backgrounds, and (where appropriate) first vaccine received.

Ethical approval was granted by the UK Health Security Agency (formerly Public Health England) Research Ethics and Governance Group (PHE REGG): Reference R&D 466.

### Data collection and analysis

Interviews were conducted via Microsoft Teams, Zoom, or phone. Topic guides were informed by existing literature, and included questions to explore experiences of the vaccine process (for those who had received one dose), beliefs about vaccines, possible reasons for not having either a vaccine, and reactions to possible government strategies aimed at improving uptake. All participants received a £40 voucher as reimbursement for their time.

Interviews were recorded with consent, transcribed, anonymised and entered into Nvivo v12. Data were analysed using a thematic approach [20, 21] and followed the process outlined by Braun and Clarke [21]. First, data were read in full by the first author (SD). Drawing on existing vaccine hesitancy frameworks we used an abductive approach to coding [22]. To begin with, codes were assigned to the data by the author without the use of pre-existing frameworks. A sample of transcripts were coded by a second author (GL) and disagreements resolved through discussion. Once data were coded in full, we drew on concepts outlined in existing frameworks; confidence, complacency, convenience, context and communication [11]. Codes were then collated into potential themes under these headings. The team met regularly to discuss the analysis process, and to review, name and define themes. The analysis continued throughout the final (writing) stage [23] with researchers exploring and presenting narratives and interpretations to address the aim of the project.

## Results

A total of 70 interviews were conducted, with 35 participants who had received one dose of the vaccine, and 35 participants who had not received any vaccine. Thirty-one participants (44%) identified as men, and 37 (56%) as women. Thirty-six participants (51%) were from minority ethnic groups (Table 1).

**Table 1:** Demographic characteristics

| N (%) | One dose: N=35 (50%) | No dose: N=35 (50%) |
| --- | --- | --- |
| Gender |  |  |
| Male | 21 (60%) | 10 (28%) |
| Female | 14 (40%) | 23 (66%) |
| Prefer not to say | 0 | 2 (6%) |
| Age |  |  |
| 18-24 | 0 | 12 (34%) |
| 25-29 | 0 | 22 (63%) |
| 30-34 | 15 (43%) | 0 |
| 35-39 | 6 (17%) | 0 |
| 40-44 | 8 (23%) | 0 |
| 45-49 | 6 (17%) | 0 |
| Prefer not to say | 0 | 1 (3%) |
| Education |  |  |
| Employed full time | 20 (57%) | 13 (37) |
| Employed part time | 4 (11%) | 9 (26%) |
| Full time education | 0 | 2 (6%) |
| Unemployed | 4 (11%) | 3 (9%) |
| Home maker | 2 (6%) | 1 (3%) |
| Self employed | 3 (9%) | 0 |
| Unable to work | 1 (3%) | 0 |
| Prefer not to say | 1 (3%) | 4 (12%) |
| Education |  |  |
| Still in full time education | 0 | 2 (6%) |
| Before finishing school | 0 | 0 |
| After finishing school | 6 (17%) | 7 (20%) |
| After finishing college | 10 (29%) | 11 (31%) |
| After finishing university | 17 (49%) | 9 (26%) |
| After postgraduate studies | 1 (3%) | 2 (6%) |
| Prefer not to say | 1 (3%) | 4 (12%) |
| Ethnicity |  |  |
| White | 14 (40%) | 18 (51%) |
| Mixed/multiple ethnic groups | 0 | 2 (6%) |
| Asian/Asian British | 5 (14%) | 5 (14%) |
| Black/African/Caribbean/Black British | 11 (31%) | 7 (20%) |
| Other ethnic group | 5 (14%) | 1 (3%) |
| Prefer not to say | 0 | 2 (6%) |
| Vaccine |  |  |
| Moderna | 7 (20%) | NA |
| Pfizer | 21 (60%) | NA |
| AstraZeneca | 7 (20%) | NA |

### 1. Perceived benefits of having a COVID-19 vaccine

Although hesitant about receiving a first or second dose of a COVID-19 vaccine, the majority of participants did not consider themselves to be “anti-vaccine,” and were usually able to recognize possible benefits of being vaccinated for themselves and for those around them. Participants who had had one dose of the vaccine discussed two possible reasons for having done so. This included to protect themselves and those around them, and to avoid social and travel restrictions and facilitate a return to normal. Participants who had not received a vaccination were able to identify similar benefits, even though they had not had a vaccine themselves:

> *“I would say that what really motivated me was the fact that without the vaccine I would have the danger of the virus. Everyone was scared of the virus. There were some talks that it is the end of the world. Everyone was scared but it was a virus that spread to every part of the world that also caused more harm and danger there. So I think the fear of getting the virus motivated me to go and get the vaccine” (Vx011, received one dose)*.

Those considering themselves to be of low risk of being seriously ill with COVID-19 often acknowledged that a possible reason for being vaccinated could be to protect others. Whilst not considering themselves to be vulnerable, these participants accepted that COVID-19 still posed a serious threat to other, more vulnerable, populations:

> *“[Vaccination is] proven to be successful when a lot of people, and the people it’s helping especially is obviously the older generation. I’m a lot less vulnerable than them, if I can help other people out by getting it. So although I said that I decided to originally not get it, that has now changed. I will get it” (Nvx034, not vaccinated)*.

In contrast to those being vaccinated to protect themselves/others from COVID-19, some participants who had received one dose of the vaccine reported having done so (at least in part) to facilitate travel and avoid restrictions being imposed on them:

> *“The place that I work – I reached a point where it was prerequisite for you to be vaccinated and to show the record, and to indicate there the evidence that you’ve been vaccinated before you are allowed to get into the job…*.*I had no other choice but to get the vaccination” (Vx020, received one dose)*.

These participants did not appear to be concerned about protecting themselves from COVID-19, and often expressed the view that they may be getting vaccinated for the ‘wrong reason’:

> *“I love my holidays so I was hearing that if you’re not vaccinated, you can’t go on holiday so that was the only reasons why I went and had it done… The only reason I would probably really have [a second dose] is because I want to go on holiday*… *that’s not really the right reasons to have it. It’s not like I think, ‘Oh, I’ve got to have it or else I might die*.*’, it’s like, ‘Oh, I really wanna go on holiday and if the government’s saying the only way I can go is if I have this frigging vaccine” (Vx008, received one dose)*.

### 2. Barriers to uptake of the vaccination

#### Perceived lack of benefit

Many participants who had not been vaccinated reported not having done so because the main reasons for being vaccinated (as described above) were not considered to be important, relevant or achievable with the current vaccine:

> *“I just feel like I don’t need it… I suppose I’ve just heard that people have said it’s not necessary, you don’t really need it, that’s what I’ve heard from other people… so I just feel like it’s not necessary for me” (Nvx010, not vaccinated)*.

In many cases, participants did not consider themselves to be at risk from COVID-19, either because they considered themselves to be young and healthy enough to fight the virus without vaccinations, or because they did not consider COVID-19 to be a threat:

> *“Yes. I don’t really understand what the vaccine is going to do. Okay. It says it’s not going to hospitalise me, but then, it didn’t do that to me in the first place, and neither did it do anything to all the other of my friends who got it at the same time as me” (Nvx032, not vaccinated)*.

> *“When they talk about the survival rate once you’ve contracted it, why are we vaccinating so much for something that survival rate’s something like 99% or 98% or something like this? That, to me, doesn’t add up” (Nvx002, not vaccinated)*.

As described above, a key motivation for having a vaccination was to keep others safe. However, participants questioned the logic of this, asking why they needed to be vaccinated themselves if vulnerable individuals are protected through their own vaccination. Participants reasoned that if a vaccine can protect the vulnerable, the “healthy” population do not need to be vaccinated, and if the vaccine is not effective enough to protect the vulnerable populations, having the (apparently ineffective) vaccination themselves also would not help:

> *“The protected, how can they need protection from the unvaccinated? That alone doesn’t make any sense. You’re either admitting that the vaccine doesn’t work or you’re basically just fully tyrannical” (Nvx029, not vaccinated)*.

### Efficacy and effectiveness

A second common reason for not being vaccinated was because participants did not think the vaccine was sufficiently effective for reducing transmission, keeping people safe, and facilitating a return to normality. In particular, many people were concerned that they would still be able to catch and transmit the virus if they were to be vaccinated:

> *“As I say, even if I had the vaccine I still might pass it on. I still might catch it. You just never know and that’s the thing” (Nvx015, not vaccinated)*.

Participants referred to the rising number of new cases of COVID-19, as well as people they knew who had caught COVID-19 after receiving both doses of the vaccination, and questioned the purpose of having a vaccine that did not prevent transmission:

> *“Well we’ve looked at figures recently and statistics that show that lots of people who have had both jabs and are both double vaccinated are still in hospital with COVID. My ((relatives)) actually, she was double vaccinated but she still got COVID and she died two week’s later. So I don’t think they can sit here and say it’s 98% or 99% effective or whatever they’ve said the stupid high percentages are, because people are still getting COVID, whether they’ve got the vaccine or not, and people are still dying” (Nvx030, not vaccinated)*.

Participants were concerned that they vaccine would not protect them against new, or future, variants, or stop them being seriously ill. Indeed, some even considered the current vaccine to be already ‘out of date’ and were not willing to take a vaccine that could not protect them against future variants:

> *“Obviously different strains are immune to the vaccine, or at least the vaccine’s not as strong against them, so I suppose I’m quite sceptical as to how the vaccines are working and whether or not it’s actually causing that much of an impact or whether it’s going to be a bit of an issue further down the line as they’ve become more and more immune” (vx018, received one dose)*.

The belief that the vaccine cannot prevent transmission resulted in participants perceiving it as ineffective for protecting those around them, and as a result, were not willing to be vaccinated to protect others:

> *“My main concern was obviously passing anything on to my wife because she still suffers with long COVID. However, having a vaccination doesn’t prevent you from getting the COVID and also it doesn’t prevent you from passing it on” (Vx009, received one dose)*.

Likewise, this belief also resulted in participants considering the vaccine insufficient to facilitate a return to normal:

> *“They sort of seem to have vaccinated 90% of the population, and now they’re talking about fire breaks in October. What was the point in that? I feel like its value has diminished somewhat. We’re going to vaccinate everyone by the end of July, and it was a really successful programme, but actually if you’ve not really achieved anything from it” (Vx003, received one dose)*.

Such concerns about the effectiveness of the vaccine resulted in some participants wondering if a “better vaccine” may increase uptake:

> *“A better vaccine I think, just a vaccine that does what I thought a vaccine did” (Nvx006, not vaccinated)*.

### Safety concerns

Concerns about the safety of the vaccine were frequently mentioned. Participants were worried about potential short and long-term side-effects, and often described concerns that there had been insufficient research conducted over an insufficient timeframe to feasibly understand the possible negative impacts of the vaccine. Participants reiterated that they were not “anti-vaccine” or “conspiracy theorists”, but had concerns and a need for information from trustworthy sources:

> *“I am not completely against and potentially in a year’s time when I know that I have taken the time to do the research and more research has been put into the vaccinations I might be more than happy to have it. Right now, at this current time I am not willing to do it quite yet” (Nvx004, not vaccinated)*.

Many participants described concerns about a range of possible side-effects; either from media, or from friends and family members who had had health problems after their vaccination:

> *“I know her boyfriend, who suffered from neurological problems and developed neurological issues where they’ve ended up in hospital, unable to walk. Just generally struggling with their mobility. Constant shakes. Not being able to look after themselves and having to have basically full-time care essentially. With no knowing if they’re ever going to fully recover…Yeah, it was all of them had it confirmed by the doctors, that it was a reaction to the vaccine” (Nvx022, not vaccinated)*.

There were particular concerns about the potential for future harms that were currently unknown:

> *“For me, it’s the long term of not knowing what it can do in the future. Although it can work now, would it have any effects on me in five years?*… *Just certain things have not been answered for me, so that’s why I’ve opted not” (Nvx025, not vaccinated)*.

Participants were particularly concerned about experiencing specific side-effects that would negatively impact something that was very important to them. In assessing the risks and benefits of the vaccination, avoidance of these “high priority” side-effects were often paramount. For example, one participant had heard that the vaccine could lead to temporary paralysis:

> *“I was like, I’m not putting myself through that. I do dancing. That would screw me. I wouldn’t be able to dance anymore if that happened to me. I didn’t want to take the risk” (Nvx022, not vaccinated)*.

Concerns about fertility and pregnancy were particularly influential among those who were pregnant or hoping to have a family in the future. Indeed, participants were often not prepared to take any risks relating to fertility or pregnancy – even if they were at an increased risk of catching COVID-19 – as the impact on their fertility or health of their child was paramount:

> *“Even if they said there was a hint of it I would probably – if it was like the tiniest, tiniest, like one in a million, that’s basically nothing, you would be very unlucky to get that one in a million*… *they’ve kind of made everyone insecure themselves by saying at the start it affects fertility” (Nvx008, not vaccinated)*.

A small number of participants had pre-existing conditions, and were concerned that the vaccine may not be safe for them; particularly if their condition had symptoms or risk factors that had been associated with the vaccine:

> *“Well basically I had a condition called Protein C deficiency which means that in situations where you can be prone to clotting like pregnancy or any long-haul flights or anything I am more prone to getting blood clots” (Nvx033, not vaccinated)*.

Participants also had concerns about the safety of the vaccines for people of different nationalities:

> *“One of my Indian friends who got vaccinated here when they went for their dose, the vaccinator told them that, ‘because you belong to a different nationality, we will keep a strict check on you because there is the possibility that you might get some serious side-effects as well’, so I was just concerned about the same” (Nvx007, not vaccinated)*.

Whilst accepting that many side-effects were unlikely, the fact that they had been mentioned was enough to cause concern:

> *“You’ve seen the online clips of people who have maybe passed away after having the vaccine, or the people who have had adverse reactions. That freaks you out, because although they say it’s one in how many that could have that effect, what if you’re that one?” (Nvx025, not vaccinated)*.

### Lack of opportunity

In the majority of cases, participants described the relative ease with which they had been able to book and attend the vaccination appointments. However, a small number of participants still reported being unable to book or attend an appointment with the ease that would be required:

> *“I am planning on getting the vaccine. But it’s like every time I think oh I’ll go and get it, everywhere’ s closed! [laughs]” (Nvx017, not vaccinated)*.

Even those who reported being highly motivated to be vaccinated to protect themselves described how important it was that vaccination appointments were accessible given that many have many other commitments:

> *“Actually I have a plan to go again but my job is really taking up my time*… *I would like to think the second dosage should be more easy to go in person… it will give me another chance that I’m free from the danger of the virus” (Vx011, received one dose)*.

As receiving the vaccination was not always a priority, vaccination appointments must be as convenient as possible:

> *“I probably will. It’s more just a matter of convenience now. If I’m free that day, then I would, but if I have anything else to do, then I wouldn’t let that get in the way” (Vx034, received one dose)*.

It was noted that centres must be easily accessible, in terms of both timing and location, if those most hesitant are to present for their vaccinations:

> *“I think if there was a local walk-in centre in our area, I think a lot more people would get it done because I think our Asian community, they are a bit wary of getting it done. Once or twice there have been people talking about the vaccine going around our local market and I have mentioned it to them to say that, if you want more people to get jabbed you need something local for them to go to” (Vx029, received one dose)*.

## 3. Assessing the risks and benefits

Whilst the barriers described above may be considered in isolation, many participants described attempts to weigh up the relative costs and benefits associated with having the vaccine; drawing on their understanding and beliefs about the efficacy and safety of the vaccine, as well as their own personal need. Participants who had had one dose of the vaccine described consideration of the various factors, and concluded that the benefits had, at the time, outweighed the possible harms:

> *“I had concerns that there was not enough time to allow for any side-effects to actually happen. I thought that the vaccine was a bit rushed into, you know, being widely available. I do understand all the steps of trialling vaccines, I do understand that measures are taken to prevent for it to have widespread serious effects on people. So on balance I do think that there are more pros than there are cons on taking the vaccine” (Vx021, received one dose)*.

In contrast, those who opted not to be vaccinated arrived at the conclusion that the benefits did not outweigh the risks:

> *“I mean I’m 24 so I think there is more, I think the risks outweigh the benefits for me in my case, because I’m not vulnerable, so for me I don’t see the benefits because I mean if there is a vaccine and you’re 100% certain that you wouldn’t catch COVID, that you can’t transmit it, then maybe, I mean I think I would probably have” (Nvx013, not vaccinated)*.

Some participants were concerned both about the risks from COVID-19 and from the risks associated with vaccination. These participants reported considerable distress in attempting to select the best option for them:

> *“I feel like I’d be an experiment and I feel like the risk, because it’s unknown… We currently don’t know what could happen or what effects it could have and I think it’s the anxiety and actually guilt – I think one of the biggest things there is guilt of, you know, I feel kind of trapped, like if I don’t do it I’m potentially going to catch COVID which would also put my child at risk, but if I do do it, I could potentially cause her to have lifelong or even fatal consequences” (Nvx002, not vaccinated)*.

There was clear evidence of uncertainty among many participants:

> *“Um, I was due to get my second dose last week and I didn’t, but I think I will get it. Um, I think I will get it but, yeah, I think, again, I kind of just need to bring myself back to that mindset of, like, it probably is better to have them both than it is to not have it at all or just have the one” (Vx007, received one dose)*.

### Factors influencing participants’ assessments of the risks and benefits of vaccinations

Factors influencing participants’ assessments of the risks and benefits of vaccination included: 1) age and health status, 2) understanding of science, 3) trust in government, and 4) pre-existing ideas and expectations.

#### Age and health status

Participants referred to their age and health status whilst attempting to assess the risks and benefits of having the vaccine. Throughout the pandemic, age groups have often been discussed, both in terms of risk from COVID-19 and their risk of side-effects from vaccines, and this was often reflected in participants’ decisions. Indeed, many viewed their risk from COVID-19 as low because they were young, fit, and healthy and did not have any underlying health conditions:

> *“My main is for the fact that I am quite young, I am fit and healthy so I feel like I could fight pretty much illnesses well on my own” (Nvx004, not vaccinated)*.

Vaccines were viewed as being necessary for elderly groups who were considered more vulnerable:

> *“My parents had [the vaccine]. They’re elderly, they’re in their mid-50s” (Nvx031, not vaccinated)*.

Due to the relatively young age of the participants, many were concerned about the longer-term impact of the vaccine on their health:

> *“As I’m quite young, I would wait at least three or four years to be sure that the side-effects are very well recognised by the doctors and the scientists, and to be sure I’m not in – it’s a big word, but danger – to have a bad side effect which could be a problem for me in my job or in my life” (Nvx018, not vaccinated)*.

The fact that participants were of childbearing age often had an impact on their decision:

> *“I am only 26 and I am looking to try to have a family of my own. When the first wave of vaccinations come out it wasn’t safe for pregnant women to have it…*.*What if I find out in five years’ time after I have tried for many, many years that I can’t conceive and it fell down to that?” (Nvx004, not vaccinated)*.

#### Understanding of science

Participants were often very keen to do their own research and to understand the role of the vaccine in the pandemic. However, information about the risks and benefits of the vaccine often caused confusion:

> *“Yeah, so the efficiency of it, I guess, because they’ve talked about sort of, oh, you’re 70%, like the number was 70% after you’d had one jab… I assumed that meant you were 70% immune from it, and then when you had the second one you were like 95 or whatever, but then they’re like, oh no, you can still transmit it and you can still catch it and go to hospital, so I’m not sure whether it’s 95% reduced symptoms? I’m not really sure what those numbers refer to anymore” (Nvx006, not vaccinated)*.

Whilst scientists themselves were usually considered trustworthy, participants often raised concerns about the quality of the science underpinning many of the recommendations. In particular, the speed at which the vaccine had been developed and tested often resulted in concerns about the rigor of the safety and efficacy testing of the vaccine, with participants claiming it to be far too soon to be making any firm conclusions:

> *“There’s not enough research. And I think how much research will go in – we’re talking years – will go in normally to figure out whether or not something’s safe. I feel like it’s just been quite rushed” (Nvx002, not vaccinated)*.

Participants were aware of debates between scientists, and were unsure what to believe:

> *“I don’t know if these facts are actually true. Like, if you look at scientists, for example, they all have different opinions and none of those opinions are factual*… *there’s quite a few debates between one scientist the University of Cambridge and another scientist from Oxford. And you’ll notice that they don’t always agree. So, when I look at these stats, I think to myself, well, is this true?” (Vx032, received one dose)*.

Participants preferred evidence that they could see, rather than what they could not:

> *“I like to see what’s in front me, and around me… So, I’m more about just physically myself seeing things, as opposed to being told it. Because can you trust it? I don’t know” (Nvx026, not vaccinated)*.

The age of the participant also appeared to have implications for who were considered trustworthy and influential sources of information, with participants prioritising parents and peers over more formal sources:

> *“Social media is where we hear a lot of the stories…Yes, that’s where we hear the majority of it, social media. The news, the young generation, they don’t necessarily listen to the news. Definitely my generation really don’t even listen to the news. It’s all on socials” (Nvx031, not vaccinated)*.

As new evidence is emerged, the changes to public health messages further reduced confidence in the vaccination program. Participants did not have the necessary information to understand why important messages had changed, and were often unable to forget previous advice, or accept that the advice would not change again:

> *“If there are good stats on that then I’ll go with it, but all I’ve heard is like I was just saying, women can’t have it then they can, pregnant women can’t but now they can, kids can’t, it needs more clarification, it needs to be more clear for people. Because that’s what’s going to put a lot of people off by just saying things like that” (Nvx008, not vaccinated)*.

Participants who had had one dose often reported feeling ‘deceived’ or that they had not been given all the information ahead of receiving the vaccination:

> *“I’d rather be given all the information and then make an informed decision, other than being given small bits of this, and then make a decision, and then get more information that then makes the previous decision that I made look a bit wonky, because I feel that they weren’t clear” (Vx031, received one dose)*.

This resulted in a situation in which many reported a preference for waiting until all the information was available:

> *“I mean I would be happy to take a vaccine once it’s not in trial anymore and that we have the full set of data” (Nvx013, not vaccinated)*.

#### (Mis)Trust in government handling of the pandemic

Many participants reported how concerns about the benefits, effectiveness and safety of vaccinations had been exacerbated by a lack of trust in the current government and their handling of the pandemic. Indeed, there appeared a generic distrust of the government, with many participants considering government actions to be motivated by political, rather than health, reasons:

> *“I don’t believe half what they [the government] say anyway so it goes in one ear and out the other. Look how often we get lied to by the government about stuff, so I just think I can’t trust a word they say” (Vx008, received one dose)*.

Constantly changing rules, guidance and information reduced trust in statements from the government, and led to concerns that the government would not follow through with any promises:

> *“I think the whole way through this pandemic the government has been saying they’ll do this if people get vaccinated, this will start to change if this percentage is hit in certain age groups, and nothing is ever followed through” (Nvx030, not vaccinated)*.

Some of the previous decisions made by the government were viewed as attempts to increase infection so that vaccinations would be necessary:

> *“One thing you can’t trust about government is they all stand clear of this section. Why did they take so long to lock down the country? Why did they take so long? Did they want more people to be infected?*… *Is that a promotion for more people to get ill and get vaccinated?” (Nvx021, not vaccinated)*.

Potential ulterior motives were suggested, such as tracking the movement of the general public, or for financial reasons:

> *“Is this all just a ploy to get us all home and under government control?” (Nvx035)*.

> *“I think [the need for a third dose is] just ridiculous. They’re just trying to get more money. They get money per vaccination…. Next thing you know it’s going to be a case of people having to get it three / four times a year, just to keep it up. I think that side of it is more just money grabbing, to be honest” (Nvx022, not vaccinated)*.

Furthermore, many organisations, such as the NHS and the BBC were considered to be under government control, resulting in a situation in which participants did not know who they could or should trust:

> *“I think all the information I’ve mainly seen is from like the BBC. So media outlets or I think NHS and stuff like that [is like] directly hearing from the government such as Boris” (Nvx027, not vaccinated)*.

> *“Um, no, I don’t think so because I like to think that I trust my midwife and my midwife’s telling me to have it, but she’s only getting her information from the government so, again, no” (Vx008, received one dose)*.

#### Pre-existing ideas and expectations

Participants’ prioritised their pre-existing ideas and experience of COVID-19, even when it conflicted with public health messages. Indeed, it was often thought that people were seeking out data that matched their existing perspective:

> *“I think if people are choosing to believe them, they use them as almost like a confirmation, ‘like oh I read…’ and it’s like, ‘yeah, but you read it on a footballer’s Twitter*.*’ No one is stupid enough to think, ‘ah, he’s stumbled upon this big conspiracy*.*’ I think people just use it as corroboration for what they already think” (Nvx006, not vaccinated)*.

Previous experience of having caught COVID-19 appeared to reduce participants’ perceptions of risk of (severe) COVID-19 infection in the future:

> *“I had COVID quite a few months ago now actually and it was definitely bad, for me it was exactly the same as having the flu, which I’ve had before, but it was never at the point where I was thinking right, I’m going to die. So I think for me, I’d much rather have my immune system fight it off naturally rather than having a vaccine that’s been generated so fast and so quick with not any long term study” (Nvx030, not vaccinated)*.

Pre-existing knowledge of certain events could reduce perceptions of how severe the virus could be:

> *“I try and look at the source or the amount of people used, if there was a study or what it was based on, the statistics, where it came from. There were some things I didn’t agree with, like, I had three friends that died from COVID, I say from COVID, it went down as COVID, none of them actually died from COVID, they had it but then had another illness after but because it was COVID within 28 days, it goes down as a COVID death, which wasn’t what caused it, they actually were doing fine with that” (Vx031, received one dose)*.

Participants’ knowledge of others having caught the virus after being vaccinated were less likely to accept that the vaccines were effective:

> *“I have a cousin who he’s been vaccinated, and he just got COVID. Apparently if you have the jabs then your symptoms might not be as bad. But then I know, I’ve heard of people who have had it since they got vaccinated, and they’ve had really bad cases. So I don’t know how true that is either” (Nvx019, not vaccinated)*.

And those with knowledge or experience of side-effects from vaccines were less willing to accept that vaccines were safe:

> *“My partner has been ill until now and that really has been what has made me rethink my position… I just thought, “What if this happens to me?” I just thought I’m going to put it on hold and I haven’t really – for me it has just been the case of I am a lot more careful, I still wear a mask everywhere I go, I go out only when it’s needed. I haven’t taken the second dose because I am scared” (Vx021, received one dose)*.

### 4. Potential impact of initiatives aiming to improve uptake on decision making

Participants discussed the potential impact of initiatives for improving uptake on their decisions. Initiatives included 1) incentives and reimbursements and 2) vaccination passes.

#### Incentives and reimbursements

Whilst the offer of financial reimbursements to cover the costs and time taken to attend vaccination appointments was viewed favorably, participants generally thought it would be preferable to make it easier for participants to be vaccinated:

> *“Frankly, I think it will be much easier if, like I said if it was easy to access places, rather than about the money. So you don’t have to take time off work” (Vx028, received one dose)*.

A small number of participants thought the offer of financial incentives (rather than reimbursements) may lead to an increase in vaccination uptake among certain populations, such as for those on a low income:

> *“Give it to those in the low-income rates. For those on high income rates they are capable of getting the vaccine” (Vx012, received one dose)*.

However, most participants thought incentive schemes were wrong; describing them as coercion, blackmail, and bribery and felt it would further increase mistrust in the vaccination program:

> *“It would not make me more likely. It would make me think, ‘Why are you bribing me to get it done?’” (Nvx025, not vaccinated)*.

Many participants thought incentivizing people would be unfair to those already vaccinated, and there were queries about where the money would come from:

> *“Where is that cost going to come from? Is it going to come out of my taxes? (Vx031, received one dose)*.

Participants who were worried about side-effects or the safety of the vaccine reported money would not alleviate their concerns:

> *“When it comes to my health, I’ve learnt that that’s something that I will not jeopardise” (Nvx027, not vaccinated)*.

Participants reported that it would take considerable sums of money to convince them to potentially risk their health; and raised concerns about the ethical issues associated with this:

> *“I mean my favourite headline was ‘kebabs for jabs’…That’s also targeting, in a way, that kind of targets people who might be in really bad financial situations and really bad poverty. Like, oh if I go and get the jab, I can eat dinner tonight. It didn’t really sit right with me to be honest” (Nvx022, not vaccinated)*.

Generally, it was thought that the money would be better spent elsewhere:

> *“I would much rather the money was spent on developing a vaccine that actually worked properly if that makes sense” (Nvx024, not vaccinated)*.

Vaccine passes

Many participants reported that the introduction of vaccination passes could increase vaccination uptake. Indeed, as described above, a number of participants described vaccination passes as being the only reason that they would have a vaccination. However, this suggestion was not well received:

> *“It wouldn’t motivate me to. It would force me to” (Nvx035, not vaccinated)*.

Almost all participants considered this to be forcing people to be vaccinated against their will, and would most certainly increase mistrust and ill feeling:

> *“I’m totally against. As I’m French I’m very connected with how it is in France and it is very concerning for me, yeah, it is. Because we cannot force people to be vaccinated, to have a medical procedure” (Nvx018, not vaccinated)*.

Participants felt that the introduction of restrictions for unvaccinated groups could reduce confidence in the government:

> *“I know I will have it in order to travel but it doesn’t make me I guess look at the government in such a positive light” (Nvx027, not vaccinated)*.

However, it was noted that the introduction of vaccination passes would not increase uptake among those with concerns about the safety of the vaccination, and may cause considerable distress among those whose employment depends on it:

> *“I have been extremely stressed. Extremely stressed. After I voiced how I feel about the potential risk, I think to be basically forced against your will, being told, ‘If you don’t, then you will lose your job’” (Nvx002, not vaccinated)*.

Those considering the vaccine to be ineffective often reported a willingness to have a vaccine should it become mandatory, although considered this to be futile. Furthermore, concerns about the efficacy of vaccinations for preventing transmission led to concerns that vaccination passes may actually increase transmission:

> *“I probably would just go, okay, I’ll crack on and have it then. But not because I think it will particularly protect me if I’m there, because apparently it won’t” (Vx003, received one dose)*.

Those most opposed to the idea of vaccination passes reported being very unlikely to have a vaccine, despite, or even because of, vaccination passes:

> *“I think it makes me a lot less likely to get a vaccine, because then it makes me more paranoid about the conspiracy theories that I hear, and well why are they forcing so many people to get it if they don’t want to get it?*… *Yeah, I wouldn’t agree with that, I think there’ll be a huge uprising if they did that” (Nvx019, not vaccinated)*.

## Discussion

### Main findings of this study

The COVID-19 pandemic remains a major threat to the health and wellbeing of the population and vaccination uptake is likely to remain a priority for the foreseeable future. Indeed, in response to considerable increase in the number of individuals who are infected with the Omicron variant of COVID-19 it is now strongly recommended that the adults in the UK receive three doses of a COVID-19 vaccine [4]. It is therefore essential that the public are supported to receive, and continue to receive, vaccines when offered so that the protection against current and future variants is as high as possible. Our research has identified motives for having a vaccine and barriers to uptake and provides important insight into the requirements for future public health campaigns targeting vaccine uptake. Research now needs to identify how to communicate risks (from COVID-19 and vaccination) and benefits (for the individual and population) so that people can make informed personal risk assessments.

Overall, participants in the current study appeared to be vaccine hesitant rather than opposed to vaccination, and many made substantial efforts to understand the benefits and possible risks associated with having the vaccine. Those unwilling to have a vaccination at present were not sufficiently persuaded that the possible risks of having the vaccine outweighed the benefits for themselves. Participants who had received one dose of the vaccination reported having done so to protect themselves or those around them, or to avoid restrictions. Among those not accepting a vaccination, it was thought that the vaccine would not, or could not, provide the anticipated benefits, and might lead to potential harms. This is very much in line with existing literature that suggests vaccine hesitancy is the result of public confidence (safety and efficacy of the vaccine), complacency (risk and severity of COVID-19), convenience (opportunity for vaccination), communications (sources of information) and context (demographic characteristics) [10, 11]. The current study suggests that these factors appear to be influenced by the age and health status of the participant [13], trust in government [15] and knowledge and understanding of scientific evidence.

Many participants in the current study considered themselves to be young and healthy, and this appeared to have a substantial impact on their decision. Throughout the pandemic, people under the age of 50 years have often been considered “low risk” from COVID-19, resulting in a situation in which participants did not consider the vaccine to be necessary to protect themselves. Whilst there was variation between participants regarding how serious COVID-19 is for others, many were concerned that the vaccine was not sufficiently effective for reducing transmission and therefore for protecting those around them. Participants were regularly exposed to reports of people who had caught COVID-19 following vaccination, either through personal connections or social media, and this often made them question how well the vaccine was working. Importantly, this “direct experience” was often given more weight than scientific evidence. Attempts to persuade young people of the benefits of vaccination may be improved through providing a clear rationale for receiving the vaccinations in light of their perceived low-risk status. Attempts to increase motivation to be vaccinated to protect others must address misconceptions regarding the impact of the vaccine on transmission of COVID-19 at a population level. However, this is not without challenges as it is also important to ensure that people know that the vaccine cannot be 100% effective so that other precautions are still taken. Indeed, it is critical that messages inform the public of the benefits and limits of the vaccine without also undermining confidence in the vaccine.

Media reports of severe side-effects among those under the age of 40 years have been prevalent throughout the rollout of the vaccine [24]. However, whilst participants were aware of possible negative reactions to the vaccine (such as blood clots), the majority of participants in the current study felt that their young age put them at greater risk of side-effects that may not yet have been identified. Indeed, it was noted that the vaccine had been developed much more quickly than other vaccines, and there were concerns that this meant it had been “rushed” and was lacking the scientific rigor and long-term follow up periods that would ordinarily be employed [25]. This concern was further exacerbated through apparent changes to recommendations, such as the apparent change in vaccination recommendations for pregnant women. Participants were aware that there were debates among scientific communities, and this further reduced confidence in the safety of the vaccine. Participants reasoned that new information about long term safety of the vaccine may come to light at any point and preferred to wait until all the information was available before receiving the vaccine. In order to reassure young people of the safety of the vaccine, it may be beneficial to emphasize what has been done and is continuing to be done to ensure safety.

In attempting to weigh the risks associated with receiving the vaccine against the risks associated with COVID-19, particular weight was given to side-effects or symptoms that were particularly important to the participant. For example, participants were of childbearing age and the potential impact of the vaccine on fertility or their unborn child was often given priority over any concerns about catching COVID-19. Importantly, these concerns remained a priority even when participants considered them to be unlikely to happen to themselves. Messages focusing on age specific concerns may provide more reassurance than those that focus on harms such as blood clots, hospital admissions or death.

Previous research has highlighted the impact of public distrust in the government on the uptake of COVID-19 vaccines [15, 16], and has highlighted the need for effective communications to address this [26]. In accordance with previous research, participants in the current study also reported a lack of confidence in the government and described how this lack of trust had informed their decision not to be vaccinated. Furthermore, key organizations such as the National Health Service (NHS), the British Broadcasting Corporation (BBC) and other media outlets were often considered to be “under government control.” As a result, participants were unclear who they could trust and what they could believe. It is therefore critical that attempts to promote vaccine uptake come from sources that are trusted and respected among young people. As social media is often a preferred source of information it is critical that attempts are made to create messages that do not become lost within the plethora of (mis)information that already exists.

Attempts to increase uptake, such as vaccination passes and financial incentives, were often considered ineffective for increasing uptake and for preventing transmission of the virus and may even be viewed as coercion. For some, this may further reduce trust in the government and science regarding the safety and efficacy of the vaccine. For those who had not been vaccinated because they did not think vaccination was necessary, the introduction of vaccination passes could provide a powerful motive. However, participants cautioned that their reason for having the vaccine would then be to avoid punishment, rather than for the benefits it provides. It was thought that this may result in negative feeling toward those mandating the vaccine. Furthermore, for participants who are concerned about the safety of the vaccine, the introduction of vaccine passes could cause considerable distress, particularly if their employment or social life is likely to be negatively impacted.

### Limitations

Despite best efforts, our recruitment strategy may have missed relevant voices, including those who are not computer literate. Our data must be interpreted with this in mind.

### Conclusions

In conclusion, the majority of young people described and concluded that for themselves, the benefits of vaccination did not outweigh the perceived risks to themselves. They did not consider themselves to be at risk of becoming seriously ill from COVID-19 and did not think that the vaccination was capable of protecting those around them. This, combined with concerns about the safety of the vaccine, resulted in reluctance to be vaccinated at present. Perceptions of risks and benefits were influenced by participants’ age and health status, trust in government, understanding of science, and pre-existing ideas and expectations. Participants were unsure who they could and could not trust and were resistant to attempts that were viewed as coercive. In order to promote uptake, public health campaigns should focus on the provision of information from trusted sources that carefully explains the benefits of vaccination and addresses safety concerns more effectively. To overcome inertia in people with low levels of motivation to be vaccinated appointments must be easily accessible (both in terms of location and timing). Research now needs to identify how to communicate risks (from COVID-19 and vaccination) and benefits (for the individual and population) so that people can make informed personal risk assessments.

## Data Availability

All data produced in the present study are available from the corresponding author upon reasonable request

## Declarations

### Ethics approval and consent to participate

Ethical approval for this study was granted by UK Health Security Agency’s (formerly Public Health England) Research Ethics and Governance Group (ref R&D 434). All participants provided informed consent to participate. All methods were carried out in accordance with relevant guidelines and regulations.

### Consent for publication

NA

### Availability of data and materials

The datasets used and/or analysed during the current study are available from the corresponding author on reasonable request.

### Authors’ contributions

Conceived the study: All authors

Study design: All authors

Collected data: SD, FM, HW, LT

Analysed the data: SD, GL, LT

Interpreted the data: All authors

Drafted the manuscript: SD

Reviewed the manuscript and approved content: All authors

Met authorship criteria: All authors

## Acknowledgements

This study was commissioned by the Department of Health and Social Care through the National Institute for Health Research (NIHR) Health Protection Research Unit (HPRU) in Behavioural Science and Evaluation.

This study is funded by the National Institute for Health Research Health Protection Research Unit in Behavioural Science and Evaluation, a partnership between the UK Health Security Agency (UKHSA) and the University of Bristol. The views expressed are those of the author(s) and not necessarily those of the NIHR, UKHSA or the Department of Health and Social Care.

SD, MH, GL, HW, RA and IO are supported by the National Institute for Health Research Health Protection Research Unit (HPRU) in Behavioural Science and Evaluation, a partnership between the UK Health Security Agency (UKHSA) and the University of Bristol.

LY is an NIHR Senior Investigator and her research programme is partly supported by NIHR Applied Research Collaboration (ARC)-West, National Institute for Health Research Health Protection Research Unit (HPRU) in Behavioural Science and Evaluation, and the NIHR Southampton Biomedical Research Centre (BRC).

FM, HW and RA are supported by the National Institute for Health Research Health Protection Research Unit (HPRU) in Emergency Preparedness and Response, a partnership between UKHSA and King’s College London.

